# Prevalence and associated factors of depression among junior healthcare professionals of public teaching hospitals of Bangladesh: An analytical cross-sectional study

**DOI:** 10.1101/2022.10.23.22281426

**Authors:** Gazi Md. Salahuddin Mamun, Md. Zakiul Hassan, Aysha Khatun, Md. Farhad Kabir, Shamsun Nahar Shaima, Sadia Afrin, Nuhad Raisa Seoty

## Abstract

Due to the significant number and its effects on quality of life, depression is becoming a major concern worldwide. Though its prevalence among junior healthcare professionals is also increasing day by day, still very few data are available regarding this. So, we’ve conducted a study to find out the prevalence and associated factors of depression among this vulnerable population. A total of 218 participants were enrolled from two public teaching tertiary-level hospitals in Dhaka, Bangladesh from October 2018 to April 2019. Data were collected by using a self-administered questionnaire including the WHO-5 well-being index. Prevalence of major depression was found at 17.9% and poor-emotional well-being was 25.2%. Factors associated independently with major depression were those thinking to be a doctor as the wrong decision (aRRR: 6.85, 95% CI: 1.40-33.45, p=0.017) and taking sedative or anxiolytic drugs (aRRR: 4.54, 95% CI: 1.50-13.73, p=0.007). On the other hand, doing physical exercise (aRRR: 0.32, 95% CI: 0.12-0.89, p=0.028) and being satisfied in their current job position (aRRR: 0.07, 95% CI: 0.02-0.29, p<0.001) had significantly less chance of being suffering of major depression. Suicidal and self-hurting ideation was also found among 23.4% of participants. If these modifiable factors can be addressed properly and by taking necessary steps against these simply identifiable factors, unwanted incidences can be prevented especially in low- and middle-income countries.

**What is already known on this topic:** Depression is common among healthcare professionals but is still neglected especially in low- and middle-income countries.

**What this study adds:** Thinking of being a doctor as the wrong decision, taking sleeping pills, not doing physical exercise, and being not satisfied in their current job position are associated with depression among junior healthcare professionals. Suicidal and self-hurting ideation were also found high among the participants.

**How this study might affect research, practice or policy:** Early identification of major depression by simple factors may help to initiate prompt strategies that will reduce the burden of depression among junior healthcare professionals and may improve the healthcare services of low- and middle-income countries.

## Introduction

Depression is a common mental disorder associated with impaired quality of life [1]. It is a growing health issue in both developed and developing countries. General unawareness at the population level, lack of training among healthcare providers, and scarcity of resources including treatment opportunities may conceal the real burden of depression in developing countries [2]. According to WHO, worldwide 5% of adults suffer from depression [3]. According to a national mental health survey in 2019, in Bangladesh, 6.7% of people suffer from depressive disorders [4]. Among the WHO’s South East Asia Region, Bangladesh ranks highest in anxiety disorders [5]. Also, suicide is the 4th leading cause of death among 15-29 years old throughout the world which is a consequence of major depression [3].

Although the physical health of medical practitioners is better than average, they are at significantly higher risk of mental illness and suicide than the general population [6]. Physicians are exposed to high levels of occupational stress resulting in suffering from some mental disorders like anxiety, depression, and occupational burnout [7]. This may lead to affect their professional performance and the quality of healthcare service provided [8-10]. Beyond the effects of depression on individuals, resident physicians’ depression was found to be associated with poor-quality patient care and increased medical errors [9]. These issues have negative effects on patients’ health and the development of the healthcare system especially in low- and middle-income countries where the healthcare system is mainly based on manpower sources [11]. As a result, concern about the psychological well-being of physicians has increased and leading to further research regarding the factors that influence the mental health of physicians. Depression has a great impact on the personal and social life of an individual. It also affects the quality of work, especially in a very sensitive profession as a physician. Internship and training periods for the post-graduation are also known to be a time of high stress. Newly appointed physicians are faced with long work hours, sleep deprivation, loss of autonomy, and extreme emotional situations during this period.

In Bangladesh, the primary workforces of a tertiary-level hospital are intern doctors, honorary medical officers (HMO), and indoor medical officers (IMO). From the very beginning of their professional life, they’ve to struggle a lot to be established in society such as post-graduation of the desired subject, achieving a secured job, maintaining happy family life, etc. These struggles may lead to depression.

On the other hand, the doctor-patient relationship is not up to a satisfactory level in Bangladesh, and depression among healthcare professionals may be one of the causes of it. Though there is no available data in Bangladesh, depression may also be responsible for some preventable medical errors in Bangladesh.

Some studies have been conducted in Bangladesh to find out the prevalence of depression among different population groups, but no study is found to evaluate the mental status of junior healthcare professionals who are directly responsible for the treatment of a majority of the population and also to gain the achievements of health sectors in Bangladesh.

## Methods and materials

### Study design and population

This study was a cross-sectional study conducted to find out the level of depression among junior healthcare professionals in two selected public teaching hospitals in Bangladesh. All the intern doctors, Honorary Medical Officers & Indoor Medical Officers of these hospitals were the study population. Junior healthcare professionals having less than 6 months of working experience were excluded from the study. For those who were busy with emergency patient management and not interested to take part in this study, data were not collected also from them.

### Study setting

This study was conducted in the following two selected public teaching hospitals in Dhaka, the capital city of Bangladesh: Dhaka Medical College Hospital (DMCH) and Sir Salimullah Medical College Mitford Hospital (SSMCMH). Both of these are tertiary-level hospitals having a capacity of 2300 beds and 800 beds respectively. But most of the time, several times more patients stay in these hospitals. Every day all types of patients from different corners of Bangladesh are reported to these hospitals to get better management. They have almost all the specialty/sub-specialty of Medical science here with almost all modern investigation facilities.

### Working definitions

Junior healthcare professionals: Doctors who are working as intern doctors or honorary medical officers or indoor medical officers were considered junior healthcare professionals in this study. These are training posts. Above these posts, the mid-level physicians are assistant registrar, registrar, resident physician or resident surgeon, etc. Senior posts included consultants and professors.

Intern doctors: After completing the 5 years course of Bachelor of Medicine and Bachelor of Surgery (MBBS), each doctor has to do 1-year training as an internship to take the registration no. from Bangladesh Medical & Dental Council (BMDC). This period is known as an internship. During this training period, intern doctors can’t take part in private practice or duty anywhere other than their assigned training hospital.

Honorary Medical Officer (HMO): After completion of the internship, a doctor has to do training for the post-graduation degree according to the specialties. This training was mostly done in government medical college hospitals without any payment. For FCPS training, this period was up to 4 years. For MD or MS training (non-residency), this training period is up to 5 years.

Indoor Medical Officers (IMO): They are government officers placed in training posts at tertiary-level hospitals for post-graduation training according to the availability of posts in the hospital.

They are paid according to the government pay scale. Usually, they have to come in this post after completing a minimum of 2 years of placement in Upazila Health Complex i.e. the primary healthcare level in Bangladesh.

Depression: According to American Psychiatric Association, Depression (major depressive disorder) is a common and serious medical illness that negatively affects one’s feelings, thinking, and acting. Based on some previous study findings, having a score ≤28 on the WHO-5 well-being questionnaire was considered a major depression in this study [12]. A score <52 on the WHO-5 well-being index is suggestive of poor emotional well-being and also is an indicator for further testing [12].

Recent life changes: Recent life changes of the participants mean that was there any significant change in their life that could affect a confounder in this study. The options used for this quarry were loss of job (self or family member), move or death of loved ones, failure in any professional examination, or any other incidents like these.

### Sample size calculation

As this is a cross-sectional study, so the sample size was calculated by the n= Z^2^pq/d^2^ equation. Prevalence of depression (p) was considered 17% in a study conducted in the neighboring country among the post-graduate resident physicians in a tertiary health care institute [13]. Considering the 95% confidence interval and the degree of accuracy required (d) as 5%, the total sample size was 217. For an equal collection of data from 2 selected hospitals, data were collected from a total of 218 participants.

### Sampling technique

Study site selection: From the list of four tertiary-level public teaching hospitals in Dhaka city, two hospitals were selected by purposive sampling.

Selection of the respondents: In selected two tertiary level hospitals, 218/2 = 109 respondents from each hospital were included by convenience type of non-probability sampling until the sample size was fulfilled. Participants were included from all departments of the hospitals.

### Data collection method and instrument

A semi-structured questionnaire was developed to collect the socio-demographic, occupation, and lifestyle-related data of the respondents. The sociodemographic and lifestyle-related variables were created by literature review. To measure the level of depression, a previously verified and published questionnaire named as WHO-5 well-being questionnaire was used. After developing the questionnaire, it was pretested among ten junior healthcare professionals similar to the study population other than the study area to identify potential problems in the questionnaire. After pre-testing, the questionnaire was finalized.

Data were collected from enrolled participants from 06 October 2018 to 20 April 2019. For the enrolment, after taking verbal permission from the on-duty mid-level physicians and proper briefing about the study to the on-duty junior physicians, written informed consents were taken from those who were interested to take part in this study. Then the self-administered questionnaire was given and explained to them properly about it. Those who were free at that time, filled it up and returned it to the investigator immediately. Those who were busy had taken the questionnaire and after filling it up returned on the following days. Investigator checked for the completeness of the questionnaires during the collection from the participants.

In this study, depression was assessed by using the WHO-5 well-being index. It is not only a well-validated measure of positive well-being, but is also used in a wide range of settings including several population groups. This questionnaire has also been shown to have good sensitivity to depression [12,14]. A difference from some other scales is that it is a positive mood scale, measuring the absence rather than the presence of negative mood in the last 14 days. In this questionnaire, on a six-point Likert scale ranging from 0 (not present) to 5 (constantly present), the degree to which specifically mentioned feelings were present in the last 14 days was recorded. Then the raw scores were multiplied by four to transform into a score from 0 (worst thinkable well-being) to 100 (best thinkable well-being). The sensitivity and specificity of the WHO-5 well-being index for depression are 93% and 83% respectively [15].

### Data management and analysis

Initially, data were checked for completeness, and correctness to exclude inconsistent data. Corrected and completed data were then entered into the SPSS for Windows (version 20.0; SPSS Inc, Chicago). All statistical analyses were done using STATA SE 13.0 (Texas, USA) software. Bivariate logistic regression analysis was done to find out the associated factors of depression and expressed as odds ratio (OR). Multinomial logistic regression analysis was done to find out the independently associated factors of depression. In the regression model, depression level by WHO-5 well-being questionnaire score was the dependent variable whereas significantly associated factors with depression (from bivariate analysis) were the independent variables. The strength of association was estimated by evaluating the relative risk ratio of multinomial logistic regression (RRR). In this study, having a p-value <0.05 was considered a statistically significant factor.

### Ethical clearance

Ethical clearance for this study was taken from the Ethical Review Committee of the State University of Bangladesh before starting the study.

## Results

During the study period, a total of 218 participants were enrolled in this study. Among them 124 participants (56.9%) were normal, 55 participants (25.2%) were in poor-emotional well-being and 39 participants (17.9%) were in major depression according to the WHO-5 well-being questionnaire. Though male and female participants were equal, 167 participants (76.6%) were below 31 years old. Nearly half of the participants were intern doctors (n= 97, 44.5%), while remaining were HMO (n=52, 23.9%) or IMO (n=69, 31.7%). Almost one-fourth of the participants (n=51, 23.4%) thought that they would be better off dead or hurting herself/himself at least once within the last 14 days. Among the participants, half of them were satisfied (n=116, 53.2%) with their job (Figure 1).

**Figure 1:**
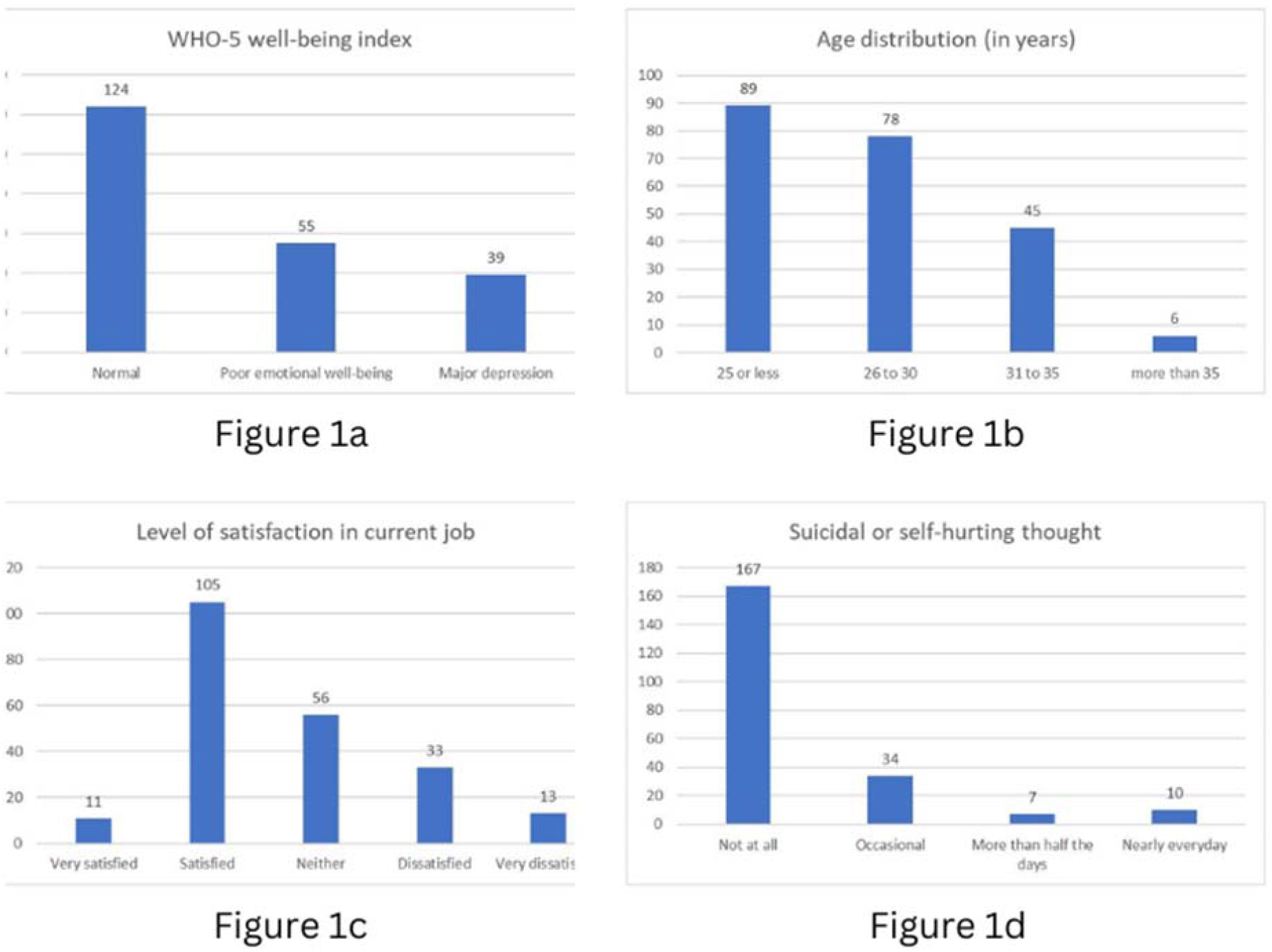
Distribution of participants by (a) WHO-5 well-being index, (b) age, (c) level of satisfaction, and (d) suicidal or self-hurting thoughts.

Compared to the participants with normal well-being, those who were in a poor emotional well-being state, came to this profession by their family choice (OR: 2.53, 95% CI: 1.32-4.88, p=0.005), thought that the decision to be a physician was wrong (OR: 5.37, 95% CI: 1.56-18.41, p=0.008) or being more confused about being in this profession (OR: 3.41, 95% CI: 1.68-6.39, p=0.001), had more chance of suffering from chronic illness (OR: 2.85, 95% CI: 1.13-7.18, p=0.026), and taking sedative or anxiolytic drugs (OR: 3.02, 95% CI: 1.39-6.56, p=0.005). Simultaneously they did not have any additional degree after graduation (OR: 0.34, 95% CI: 0.12-0.94, p=0.038), and were less likely to be satisfied in their current job position (OR: 0.27, 95% CI: 0.14-0.53, p<0.001). Other variables were comparable between these two groups (Table 1).

**Table 1:**
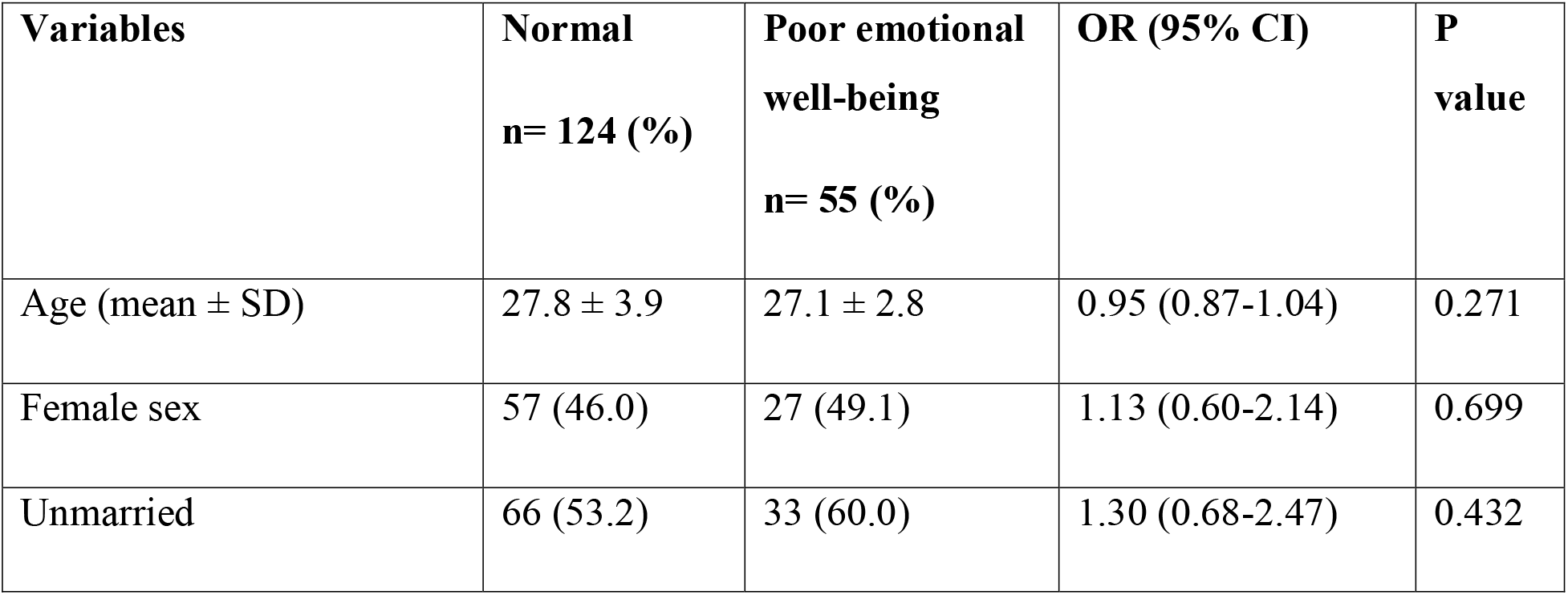
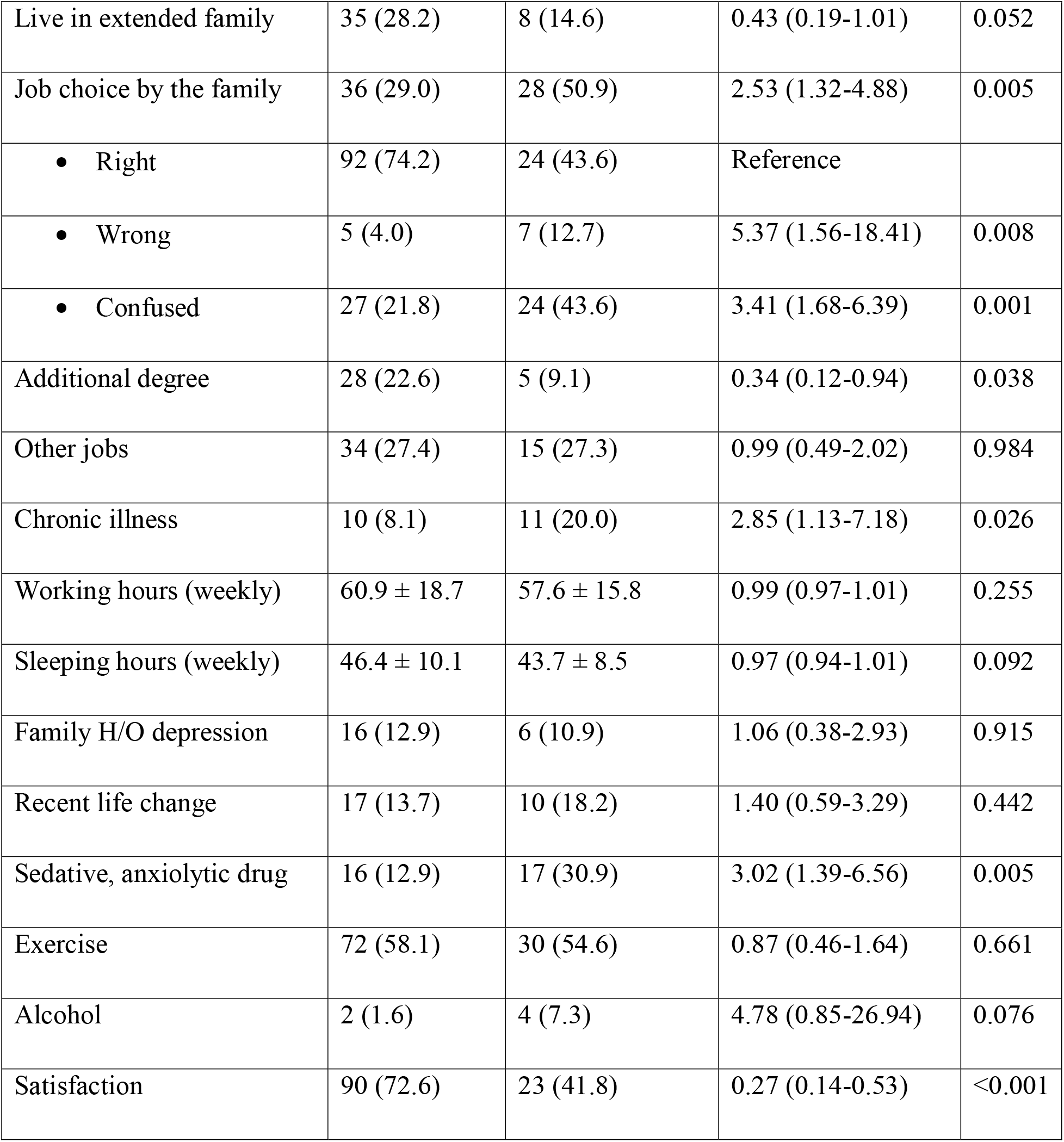
Comparison between the participants with normal and poor emotional well-being.

Compared to the participants with normal well-being, those who were in major depression, came to this profession by their family choice (OR: 3.51, 95% CI: 1.67-7.41, p=0.001), and thought that the decision of being a physician was wrong (OR: 32.71, 95% CI: 9.70-110.28, p<0.001) or being more confused about being in this profession (OR: 5.30, 95% CI: 2.07-13.58, p=0.001), had more chance of suffering from chronic illness (OR: 3.93, 95% CI: 1.50-10.34, p=0.006), family history of depression (OR: 3.37, 95% CI: 1.35-8.38, p=0.009), and taking sedative or anxiolytic drugs (OR: 9.70, 95% CI: 4.25-22.17, p<0.001). Simultaneously they had fewer sleeping hours (OR: 0.95, 95% CI: 0.91-0.99, p=0.026), were less involved in doing physical exercise (OR: 0.32, 95% CI: 0.15-0.69, p=0.004), and were less likely to be satisfied in their current job position (OR: 0.03, 95% CI: 0.01-0.11, p<0.001). Other variables were comparable between these two groups (Table 2).

**Table 2:**
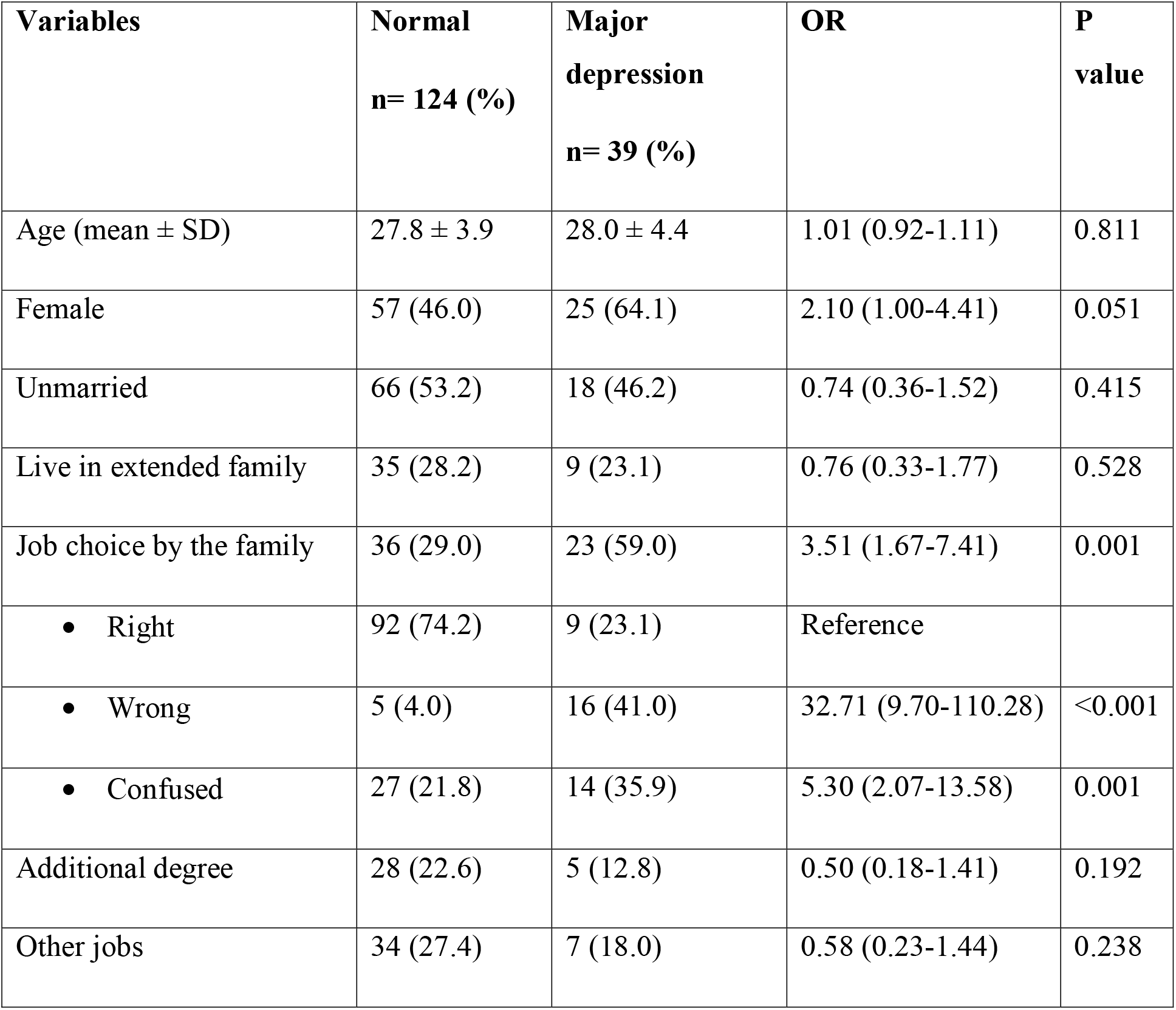
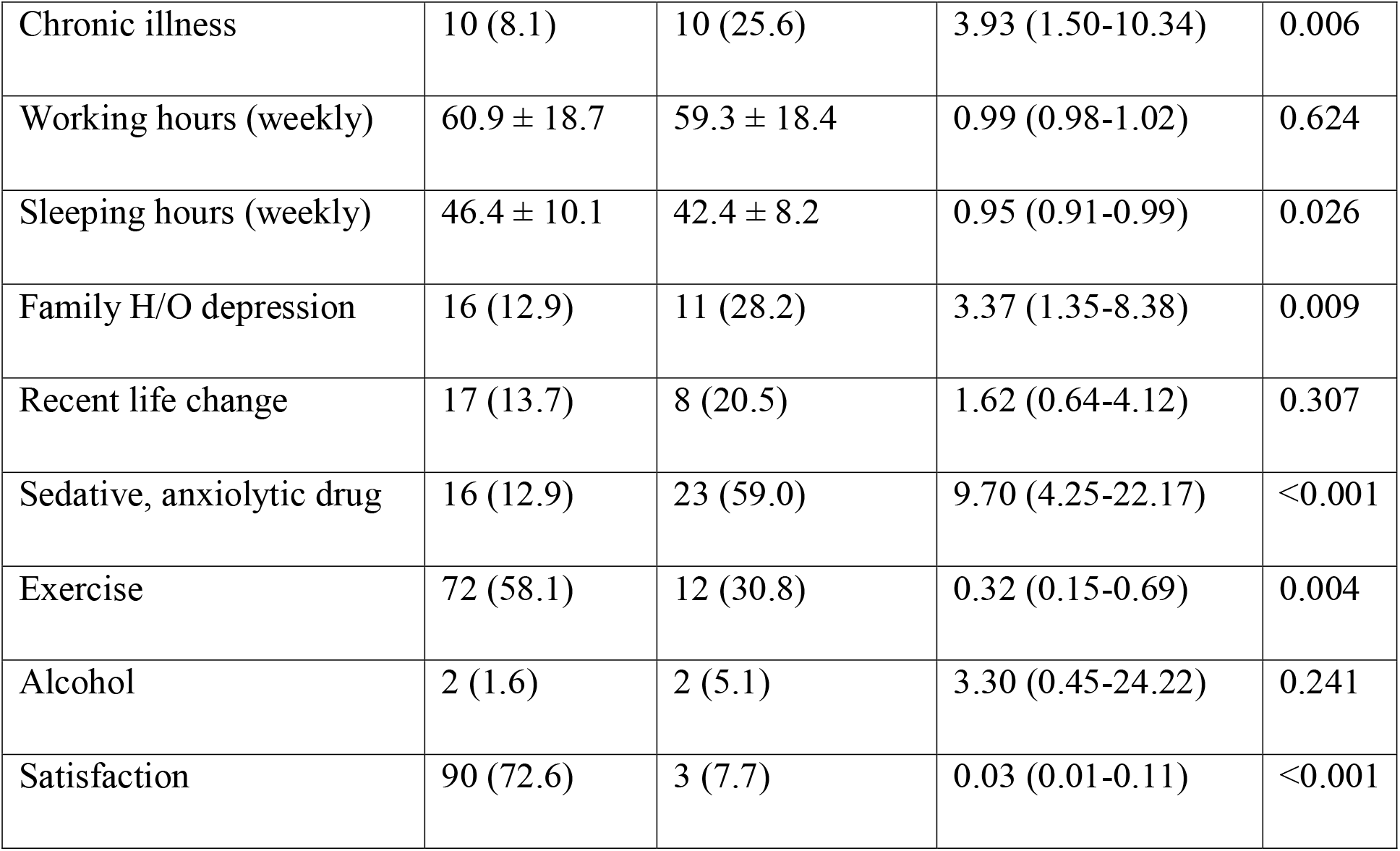
Comparison between the participants of normal and major depression.

| **Variables** | **Normal**<br>**n= 124 (%)** | **Major**<br>**depression**<br>**n= 39 (%)** | **OR** | **P**<br>**value** |
| --- | --- | --- | --- | --- |
| Age (mean ± SD) | 27.8 ± 3.9 | 28.0 ± 4.4 | 1.01 (0.92-1.11) | 0.811 |
| Female | 57 (46.0) | 25 (64.1) | 2.10 (1.00-4.41) | 0.051 |
| Unmarried | 66 (53.2) | 18 (46.2) | 0.74 (0.36-1.52) | 0.415 |
| Live in extended family | 35 (28.2) | 9 (23.1) | 0.76 (0.33-1.77) | 0.528 |
| Job choice by the family | 36 (29.0) | 23 (59.0) | 3.51 (1.67-7.41) | 0.001 |
| • Right | 92 (74.2) | 9 (23.1) | Reference |  |
| • Wrong | 5 (4.0) | 16 (41.0) | 32.71 (9.70-110.28) | <0.001 |
| • Confused | 27 (21.8) | 14 (35.9) | 5.30 (2.07-13.58) | 0.001 |
| Additional degree | 28 (22.6) | 5 (12.8) | 0.50 (0.18-1.41) | 0.192 |
| Other jobs | 34 (27.4) | 7 (18.0) | 0.58 (0.23-1.44) | 0.238 |
| Chronic illness | 10 (8.1) | 10 (25.6) | 3.93 (1.50-10.34) | 0.006 |
| Working hours (weekly) | 60.9 ± 18.7 | 59.3 ± 18.4 | 0.99 (0.98-1.02) | 0.624 |
| Sleeping hours (weekly) | 46.4 ± 10.1 | 42.4 ± 8.2 | 0.95 (0.91-0.99) | 0.026 |
| Family H/O depression | 16 (12.9) | 11 (28.2) | 3.37 (1.35-8.38) | 0.009 |
| Recent life change | 17 (13.7) | 8 (20.5) | 1.62 (0.64-4.12) | 0.307 |
| Sedative, anxiolytic drug | 16 (12.9) | 23 (59.0) | 9.70 (4.25-22.17) | <0.001 |
| Exercise | 72 (58.1) | 12 (30.8) | 0.32 (0.15-0.69) | 0.004 |
| Alcohol | 2 (1.6) | 2 (5.1) | 3.30 (0.45-24.22) | 0.241 |
| Satisfaction | 90 (72.6) | 3 (7.7) | 0.03 (0.01-0.11) | <0.001 |

After adjusting the potential confounders, in multinomial multivariable logistic regression analysis, no significant associated factor was found in suffering from poor emotional well-being compared to normal well-being status. But thinking to be a doctor as the wrong decision (aRRR: 6.85, 95% CI: 1.40-33.45, p=0.017), and taking sedative or anxiolytic drugs (aRRR: 4.54, 95% CI: 1.50-13.73, p=0.007) were significantly associated with major depression. On the other hand, doing physical exercise (aRRR: 0.32, 95% CI: 0.12-0.89, p=0.028) and being satisfied in their current job position (aRRR: 0.07, 95% CI: 0.02-0.29, p<0.001) had significantly less chance of being suffering of major depression (Table 3).

**Table 3:** Results of multinomial multivariable logistic regression analysis to explore the independent factors of poor emotional well-being and major depression.

| Variables | Normal vs Poor emotional well-being | Normal vs Major depression |
| --- | --- | --- |

|  | **aRRR (95% CI)** | **P value** | **aRRR (95% CI)** | **P value** |
| --- | --- | --- | --- | --- |
| Female sex | 0.97 (0.45-2.08) | 0.930 | 2.07 (0.73-5.86) | 0.171 |
| Extended family | 0.43 (0.16-1.17) | 0.099 | 0.73 (0.22-2.46) | 0.615 |
| Choice by family | 1.52 (0.70-3.30) | 0.291 | 1.10 (0.38-3.19) | 0.868 |
| The decision of being a doctor |  |  |  |  |
| • Wrong | 2.73 (0.65-11.54) | 0.172 | 6.85 (1.40-33.45) | 0.017 |
| • Confused | 1.80 (0.78-4.17) | 0.171 | 2.36 (0.70-7.93) | 0.165 |
| Additional degree | 0.40 (0.12-1.31) | 0.131 | 0.61 (0.15-2.51) | 0.491 |
| Chronic illness | 3.04 (0.99-9.31) | 0.051 | 3.59 (0.93-13.85) | 0.063 |
| Sleeping hours (weekly) | 0.99 (0.95-1.03) | 0.671 | 1.00 (0.94-1.06) | 0.974 |
| Family H/O depression | 0.73 (0.22-2.44) | 0.607 | 1.87 (0.48-7.23) | 0.365 |
| Sedative, anxiolytic drug | 1.84 (0.73-4.61) | 0.196 | 4.54 (1.50-13.73) | 0.007 |
| Exercise | 0.88 (0.43-1.83) | 0.740 | 0.32 (0.12-0.89) | 0.028 |
| Satisfaction | 0.46 (0.21-1.01) | 0.052 | 0.07 (0.02-0.29) | <0.001 |

## Discussion

To our knowledge, this is the first study that evaluated the prevalence and associated factors of depression among junior healthcare professionals in public teaching hospitals in Dhaka, the capital of Bangladesh. The main observation of this study is that the prevalence of major depression was 17.9% among junior healthcare professionals and was associated with thinking of being a doctor as a wrong decision, taking sedative or anxiolytic drugs, not doing physical exercise, and not being satisfied with the current job position. Besides these, 25.2% of participants were in poor emotional well-being which was associated with suffering from chronic illness, and no satisfaction in the job. Also, among the participants, 23.4% thought that they would be better off dead or hurting herself/himself at least once within the last 14 days.

Medical practitioners at all stages of their careers have higher levels of depression than the general population. A series of recent meta-analyses estimates the prevalence of depression to be 27% in medical students [16] and 29% in resident physicians [17]. Perpetuating the problem, medical practitioners may be reluctant to seek help for their own mental health issues. Whilst this is likely due to a range of factors, stigma in particular acts as a barrier to seeking appropriate care [6]. A study conducted by Grover S et al, among medical professionals of a tertiary care hospital in North India, has found that about 30.1% of physicians had depression [18]. Previous investigations from US, Britain, and Japan concluded that the prevalence of depressive symptoms among physicians varied from 10% to 15% [7]. Another study from the Netherlands indicated that depressive symptoms and anxiety were prevalent in 29% and 24% of physicians, respectively [19].

We’ve found that major depression is also higher among those who used sedatives and/or anxiolytics. Studies have shown that patients suffering from depression may present with sleep deprivation [20] and also sleep disturbance precedes depression in elderly patients [21]. Recent studies also suggest that the relationship between sleep and depression may be bidirectional [22]. So, the association of using sleeping pills may be a cause of major depression in this study.

Several studies have found that physical exercise can reduce depression and can even treat depression effectively as antidepressants [23]. A randomized clinical trial by Hidalgo et al among older adults with mild to moderate depression has also found an initial similar improvement between a supervised physical exercise program and antidepressant treatment [24]. Sometimes exercise was also suggested to people with depressive symptoms who met the diagnostic criteria for depression [25]. In our study, we’ve also found similar findings that major depression is less likely among junior healthcare providers who did physical exercise regularly.

Our study has also found a significant association between major depression with less satisfaction on the job. Studies have found a strong correlation between the inadequacy of job satisfaction with depression among physicians [26,27]. A randomized controlled trial among nurses by Ghawadra et al has shown that mindfulness-based training is effective in improving anxiety and job satisfaction [28].

Those who thought of being wrong to be a physician were more likely to suffer from major depression. Though no other study was found to evaluate this factor, this may be due to the persistent dilemma, hesitation, and due to unexpected experiences throughout the journey of being a doctor.

A study conducted by Landrigan CP et al among interns has shown that the working conditions of healthcare providers including sleep deprivation [11] affect depression. Another study by Rothberg MB et al among nurses has shown that overwork [29] can also contribute substantively to this problem. But we’ve not found any association between these two factors among junior physicians.

## Conclusions

The prevalence of depression among junior healthcare professionals including suicidal and self-hurting ideation was found to be high. Major depression was also independently associated with some important factors named as lack of physical exercise, job dissatisfaction, wrong ideation of being in this profession, and a history of using sleeping pills. Thus, the identification of these simple preventable factors may help to initiate prompt strategies that may help to reduce the burden of depression among junior healthcare professionals and may improve the healthcare services of low- and middle-income countries like Bangladesh.

## Strength and Limitation

In this study WHO-5 well-being index was used as it is a well-validated measure of positive well-being, widely used in a range of settings, and has been shown to have good sensitivity to depressive symptoms or depressive affect. Also, there was no missing data in this study as the investigator checked all of the responses during the collection from respondents. But for the same reason, as there was a social taboo about mental sickness, so the possibility of hiding some information can’t be ruled out.

## Informed consent statement

After proper counseling and informing about the study procedures, informed written consent was taken from the participants before data collection. All data were de-identified before analysis for non-disclosure of personal information.

## Supporting information

STROBE checklist

## Data Availability

All data produced in the present study are available upon reasonable request to the corresponding author.

## Data availability stateme

Data will be available on request.

Funding and Acknowledgement

There was no funding for conducting this study. We gratefully acknowledge and express our sincere thanks to all participants, and also to those colleagues who helped the investigator with their invaluable support and contribution to this research.

## Authors Contributions

Conceptualization: GMSM, NRS

Data curation: GMSM

Formal analysis: GMSM, MFK

Investigation: GMSM, AK

Methodology: GMSM, NRS

Project administration: GMSM

Resources: GMSM

Software: GMSM

Supervision: NRS

Validation: GMSM, NRS, MZH, SA

Visualization: GMSM

Writing-original draft: GMSM

Writing-review and editing: GMSM, MZH, AK, MFK, SNS, SA, NRS

All authors have read and agreed to the published version of the manuscript.

## Biography of the first author

Dr. Gazi Md. Salahuddin Mamun is a clinical researcher and mainly works on hospital-based research projects. After achieving his MBBS degree, he obtained his master of Public Health with a major in Epidemiology. He was involved in various research projects since 2016 including multi-country cohort projects on post-discharge mortality, childhood TB, randomized controlled trial among malnourished children, evaluating the use of wearable biosensor devices among septic children and adolescents, a phase-II trial of low-cost bubble CPAP device among adults, and also cross-sectional study among COVID-19 patients. Currently, he is working at International Centre for Diarrhoeal Disease Research, Bangladesh (icddr,b) for more than 8 years. His research interests include infectious diseases such as diarrhoea, pneumonia, sepsis, tuberculosis, etc., and also childhood malnutrition and mental health.

